# XPRS: A Tool for Interpretable and Explainable Polygenic Risk Score

**DOI:** 10.1101/2024.10.24.24316050

**Authors:** Na Yeon Kim, Seunggeun Lee

**Author notes:** **Corresponding author**, Seunggeun Lee, Graduate School of Data Science, Seoul National University, Seoul, South Korea.

## Abstract

The polygenic risk score (PRS) is an important method for assessing genetic susceptibility to diseases; however, its clinical utility is limited by a lack of interpretability tools. To address this problem, we introduce eXplainable PRS (XPRS), an interpretation and visualization tool that decomposes PRSs into genes/regions and single nucleotide polymorphism (SNP) contribution scores via Shapley additive explanations (SHAPs), which provide insights into specific genes and SNPs that significantly contribute to the PRS of an individual. This software features a multilevel visualization approach, including Manhattan plots, LocusZoom-like plots and tables at the population and individual levels, to highlight important genes and SNPs. By implementing with a user-friendly web interface, XPRS allows for straightforward data input and interpretation. By bridging the gap between complex genetic data and actionable clinical insights, XPRS can improve communication between clinicians and patients.

## Introduction

The polygenic risk score (PRS) summarizes the genetic contribution to complex traits by calculating a weighted sum of risk alleles that an individual possesses. The PRS has recently emerged as a promising tool for improving human health by providing insights into the susceptibility of an individual to various diseases[1-4]. Numerous PRS construction methods, including P+T[5], LDPred[6], PRS-CS[7], BayesR[8] and MegaPRS[9], have been developed. Additionally, recent advancements in cross-ancestry methods[10],[11],[12] and pseudo-R-based evaluation approaches[13] have further improved the utility of PRSs.

Explainability is crucial for the application of machine learning models, and this should be equally true for PRSs. Explaining which factors contribute to the predicted risk can increase the reliability of the predictions and help users trust the machine learning system[14-16]. In addition, this approach can facilitate communication between key stakeholders, including clinicians and patients. In this context, explainable artificial intelligence (XAI) methods have been extensively developed. However, no specific methods or tools are designed for the PRS, which can limit its application in clinical settings. The development of XAI tools tailored for PRSs is essential to increase their effectiveness.

We introduce eXplainable PRS (XPRS), a software designed to increase the explainability of PRSs by decomposing them into genes/regions and variant contribution scores. Although the PRS model is typically built as a linear model, in which the weights for each genetic variant are known, interpreting PRSs remains challenging owing to the involvement of hundreds of thousands or even millions of genetic variants. The challenge is further compounded by the notion that the functions of most genetic variants remain unknown, making variant-level interpretation particularly difficult. XPRS addresses this by mapping variants to genes or regions.

Since genes are fundamental biological units, this gene-level approach increases the interpretability of PRSs. XPRS calculates the attributed value of each gene or region via Shapley additive explanations (SHAPs), offering detailed insights into which genes significantly contribute to the PRS of an individual. Additionally, our approach assesses the contribution of each gene at the population level, further aiding in the overall interpretation of the PRS model.

Visualization is a critical component of explainability. XPRS incorporates a multilevel visualization approach. At the population level, Manhattan plots and tables highlight important genes in the PRS on the basis of the highest variance in gene contribution scores. At the individual level, XPRS visualizes attributed values of genes to pinpoint risk genes that drive the PRS value for a given individual. Additionally, XPRS employs LocusZoom-like plots to show which genetic variant influences these genes, providing a detailed view of the genetic factors contributing to an individual’s risk profile.

## Methods

Figure 1. shows an overview of XPRS, which consists of mandatory inputs and three main processes.

**Figure 1.**
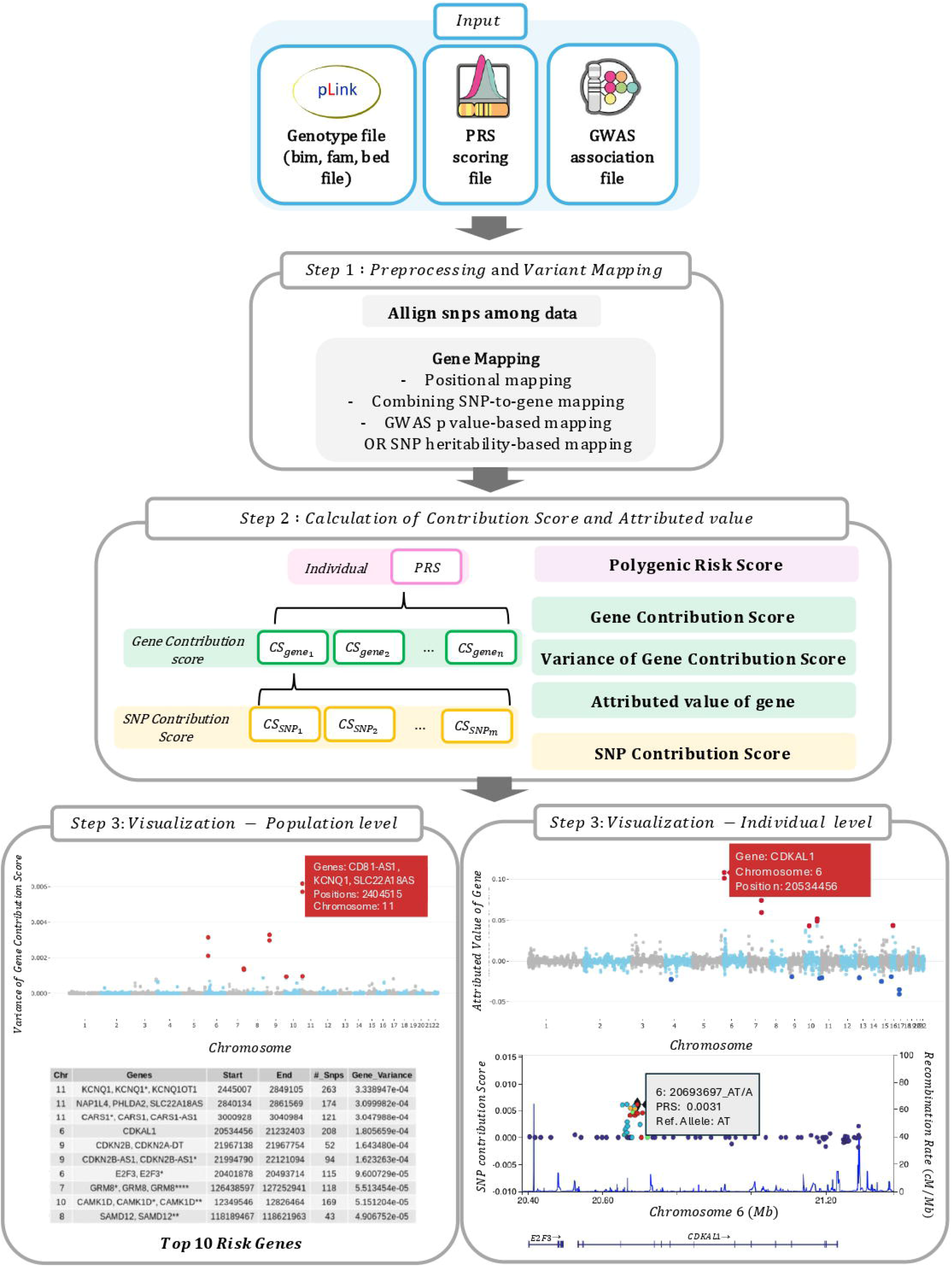
Overview of eXplainable PRS (XPRS). This figure provides an overview of XPRS. **Input**: Genotype files, PRS scoring files, and GWAS association files. **Step 1: Preprocessing and variant mapping**: SNPs are aligned and mapped to genes via positional mapping, combined with SNP-to-gene mapping, and GWAS p value-or SNP heritability-based mapping. **Step 2: Calculation**: After mapping, the polygenic risk score (PRS), gene contribution score (CS_gene_), variance of the gene contribution score, attributed value of the gene (A_gene_), and SNP contribution score (CS_SNP_) are computed in this regionizing step. **Step 3: Visualization. Population Level**: Manhattan plot of significant risk genes on the basis of variance in gene contribution scores. **Individual Level**: Density plots, gene-based Manhattan plots, and LocusZoom-like plots of the PRSs of individuals and their specific genetic contributions.

### Inputs

Our software, XPRS, requires the upload of three mandatory input files: a genotype file, a PRS scoring file, and a GWAS association file. The genotype file must be in binary PLINK format. The PRS scoring file, which is necessary for calculating the gene and single nucleotide polymorphism (SNP) contribution scores, contains information on SNPs, including SNP IDs, effect alleles, and beta coefficients corresponding to specific diseases. The file is available for download from the PGS catalog (https://www.pgscatalog.org/) or can be generated via GWAS summary statistics and reference files with various PRS construction methods. If users generate the PRS scoring file via PRS construction methods, they can optionally use GWAS summary statistics to incorporate GWAS p values for more accurate mapping information.

The GWAS association files contain a curated list of genes that have shown significant associations with specific diseases in published GWASs. Using the GWAS association file enables our analysis to focus selectively on significant genes, thereby streamlining the interpretative process by filtering out genes that do not meet established significance thresholds. At the population level of visualization, the GWAS association file ensures that only significant genes are shown, instead of all genes in a region, providing a clearer representation of risk genes. The concept of ‘regions’ will be explained in the subsequent steps of our methodology. This file can be downloaded from the GWAS Catalog (https://www.ebi.ac.uk/gwas/).

### Parameters

XPRS allows the modification of three key parameters to optimize computational efficiency and accuracy:

- CPU nodes: Users can adjust the number of CPU nodes, with a default setting of 8. When SNPs are mapped to genes, we use the parallel package in R for parallel computing to achieve faster computation.
- Top SNP heritability percentage: Users can specify that the percentage of SNPs with the highest heritability should be included in the analysis. This selection is crucial for computational efficiency and noise reduction. By default, if the number of SNPs is less than 100,000, all the SNPs are included. If it exceeds this number, users can determine the optimal percentage of top SNPs to include, whereas the default is 50% inclusion. If the default 50% inclusion results in fewer than 100,000 SNPs, XPRS automatically includes a minimum of 100,000 SNPs to ensure sufficient coverage.
- Window size: The default genomic window size is 200 kb, which is used when SNPs are mapped to genes through positional mapping on the basis of an annotation file.

These parameters increase the flexibility and performance of XPRS, allowing for efficient computation and improved interpretation of risk genes and SNPs.

### Three main processes

XPRS consists of three main steps (Figure 1). The first step includes aligning SNPs across data sources, mapping them to corresponding genes, standardizing the PRS, and adjusting beta coefficients accordingly. The second step involves calculating the gene contribution score, the attributed value of genes, and the SNP contribution score to accurately determine which genes or SNPs contribute to an increased PRS. The final step involves visualization to elucidate disease susceptibility at both the individual level and population level.

### Step 1: Preprocessing and Variant Mapping

Initial preprocessing involves SNP alignment between the genotype file and the PRS scoring file to ensure the correct inclusion of variants in the PRS. This process includes making all beta values positive and correspondingly switching the A1 and A2 allele positions to ensure that the effect sizes are consistently oriented. The raw PRS is calculated as a weighted sum of risk alleles via positive beta coefficients. The PRS is then standardized to achieve a mean of zero and a standard deviation of one. The beta coefficients are subsequently adjusted on the basis of the standardized PRS to ensure that the contribution scores derived in subsequent steps accurately reflect the standardized risk contributions. This step is followed by a three-phase gene mapping protocol:

#### 1. Positional mapping

We mapped SNPs to genes on the basis of their genomic position via refGene annotation (https://hgdownload.soe.ucsc.edu/goldenPath/hg38/database/). In cases where a gene exhibits multiple start and end positions owing to alternative splicing or isoforms, distinct start □ end coordinates are denoted with asterisks (e.g., Gene*, Gene**, and Gene***).

#### 2. Combining SNP-to-Gene (cS2G) mapping[17]

This employs several linkage methodologies, including expression quantitative trait loci (eQTL) analysis, enhancer □ gene interactions, and promoter capture Hi-C (PCHI-C) techniques, to map SNPs to genes. The cS2G file used for this step was downloaded from https://zenodo.org/records/6354007.

#### 3. GWAS p value or SNP heritability-based mapping

For SNPs not mapped in the previous two steps, we use GWAS p value significance or SNP heritability estimates. When GWAS summary statistics are provided, we identify the index SNP with the lowest p value first, assigning neighboring SNPs to the index SNP to form a region within a predefined genomic window, typically 500 kb, and continue this process until all SNPs are mapped. If GWAS summary statistics are not available, we calculate SNP heritability via the equation *heritability = β*^2^ × *MAF* × (*1* − *MAF)*, where β is the effect size from the PRS model and MAF is the minor allele frequency. The SNPs with the highest heritability are mapped first, following the same regional assignment process until all the SNPs are mapped.

Following the mapping protocol, the analysis incorporates a ‘regionizing’ step to increase computational efficiency and focus on significant risk genes. This process consolidates genes with overlapping SNP profiles into distinct regions on the basis of shared SNP content. For example, genes with identical SNP compositions are combined and calculated as single entities. In cases in which genes share a significant proportion of SNPs (e.g., two-thirds overlap), they are grouped into the same region. Within each region, we identified the gene with the highest variance in the gene contribution score to represent the risk gene. This clustering allows us to highlight the most influential genes, ensuring a more accurate and specific risk assessment by reducing redundancy and focusing on the top risk genes.

### Step 2: Calculation of the contribution score and attributed value

In step 2, we decompose the PRS into finer components for increased interpretability: gene contribution scores (*CS*_*gene*_), SNP contribution scores (*CS*_*SNP*_), and the attributed values of genes.

### Gene Contribution Score (CS_gene_) and its variance

This score quantifies how much each gene contributes to increasing the individual PRS and is calculated by summing the weighted risk alleles mapped to genes:

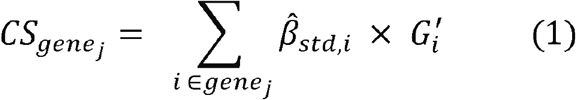

*where* 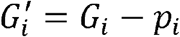

where 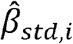 is the standardized effect size (i.e., weight) of SNP *i, G*_*i*_ is the individual genotype for SNP *i*, and *p*_*i*_ is the allele frequency of SNP *i*. Since the adjusted beta coefficients are used, the genotype data are recalculated by subtracting the allele frequency. The gene contribution score reflects how much each gene contributes to the overall PRS, providing a measure of the effect of each gene on disease risk. Additionally, we calculate the variance of 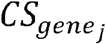 across the population to identify genes that significantly contribute to the PRS among the population. Note that the variance of 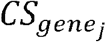 is equivalent to the heritability due to gene j.

### Attributed Value of Genes (A_gene_)

This value identifies genes that increase disease risk by comparing the contribution score of a gene to the average contribution score in the population:

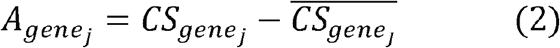

where 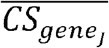 is the mean contribution score of 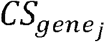 across all individuals in the population.

The attributed value highlights genes with higher-than-average contributions to the PRS, revealing those that significantly elevate disease risk within individuals. We note that 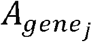 is the SHAP value in a linear model.

### SNP Contribution Score (CS_snp_)

This score quantifies the contribution of each SNP to the PRS of an individual. It is calculated by multiplying the effect size 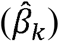 and the number of risk alleles (*X*_*k*_) that an individual carries for an SNP:

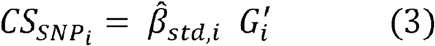

These calculations provide a detailed understanding of the PRS model. The gene contribution score quantifies the overall impact of each gene on the PRS, whereas the attributed value distinguishes genes with significant contributions compared with the population average, and the SNP contribution score breaks down the gene impact into specific SNP effects.

### Step 3: Visualization

The derived metrics from the PRS calculations are presented through a multilevel visualization approach, including both the population level and the individual level. At the population level, Manhattan plots and tables are used to identify important genes on the basis of the highest variance of *CS*_*gene*_., which highlights which genes or regions drive the PRS model. At the individual level, visualization includes Manhattan and LocusZoom-like plots, which show which regions and SNPs drive high or low values of the PRS score of an individual.

### Implementation

The XPRS software is designed to be both user friendly and efficient. We developed a web interface using Flask, enabling users to input data easily through a web page. The gene contribution scores are calculated using C++ for optimal performance, whereas R is employed for data preprocessing and visualization. This integrated approach ensures accessible and efficient handling of complex computational tasks.

### Execution

The XPRS platform was developed via a web interface with Flask, a lightweight WSGI web application framework in Python. To start the XPRS platform, the following environment variables need to be set:

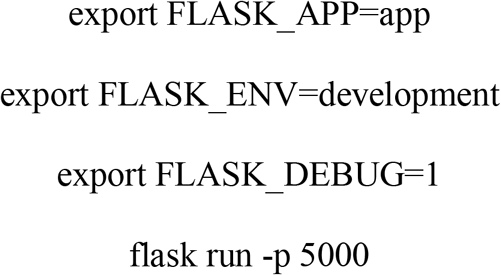

After executing these commands, a web browser will automatically open, launching the XPRS interface, allowing users to interact with the platform easily, similar to any standard web application.

### Computational Resources

When the XPRS platform runs on an AMD EPYC 7542 CPU with 8 cores, it takes approximately 5.72 minutes to visualize the contribution of genes to the PRSs for 503 samples with 80,855,722 variants in a population-level analysis. The visualization of the impact of genes and SNPs for an individual is completed in approximately 2.54 seconds.

## Results

### Web Interface

XPRS comprises three main sections accessible from the left panel: Home, Tutorial, and Run. The “Home” section provides a brief introduction to XPRS. The “Tutorial” section offers an overview of the software and the input format to ensure that users can easily understand the required data formats. In the “Run” section, there are four tabs for different genotype data input cases.

Case 1 considers a scenario in which users have a large cohort genotype file, which can be used for both population reference genotypes and genotypes of each individual for PRS prediction. With the cohort genotype file, XPRS highlights risk genes in the population. Users can conduct individual-level analysis by entering the individual ID (iid). The population-level results are saved as ‘data.rds’, enabling users to rerun analyses via previously processed data by selecting Case 1-1 and uploading ‘data.rds’, which reduces the time required for analysis.

For users without a cohort genotype file for reference but with individual genotype files only, Case 2 allows the use of reference files from projects such as the 1000 Genomes Project. This setup allows individuals to upload their genotype data and obtain personalized results and explanations. Like Case 1-1, Case 2-1 also allows users to upload the ‘data.rds’ file from the initial analysis, which is saved in the output file, to rerun the analysis efficiently.

### Visualizing the contribution of genes or regions to the PRS in the population

To identify which genes contribute the most to the PRS in a population, we visualized the variance in the gene contribution scores. The input data for this analysis included genotype files from the 1000 Genomes Project[18], with 503 samples and 80,855,722 variants from the East Asian population. The PRS scoring file was obtained from the study by Kim et al. (2024), which evaluated the PRS for type 2 diabetes mellitus in the Korean population[19]. Additionally, the GWAS association file was obtained from the GWAS Catalog (https://www.ebi.ac.uk/gwas/) for type 2 diabetes.

Figure 2 presents a Manhattan plot and a table based on the analysis. The Manhattan plot highlights the importance of risk genes associated with type 2 diabetes, where each point represents a gene and its variance in gene contribution score—higher variance indicates greater importance as a risk factor. The accompanying table lists significant genes, the number of mapped SNPs per gene, and their respective variances. Notably, KCNQ1 and KCNQ1OT1 show the greatest variances, underscoring their significant role in susceptibility to type 2 diabetes.

**Figure 2.**
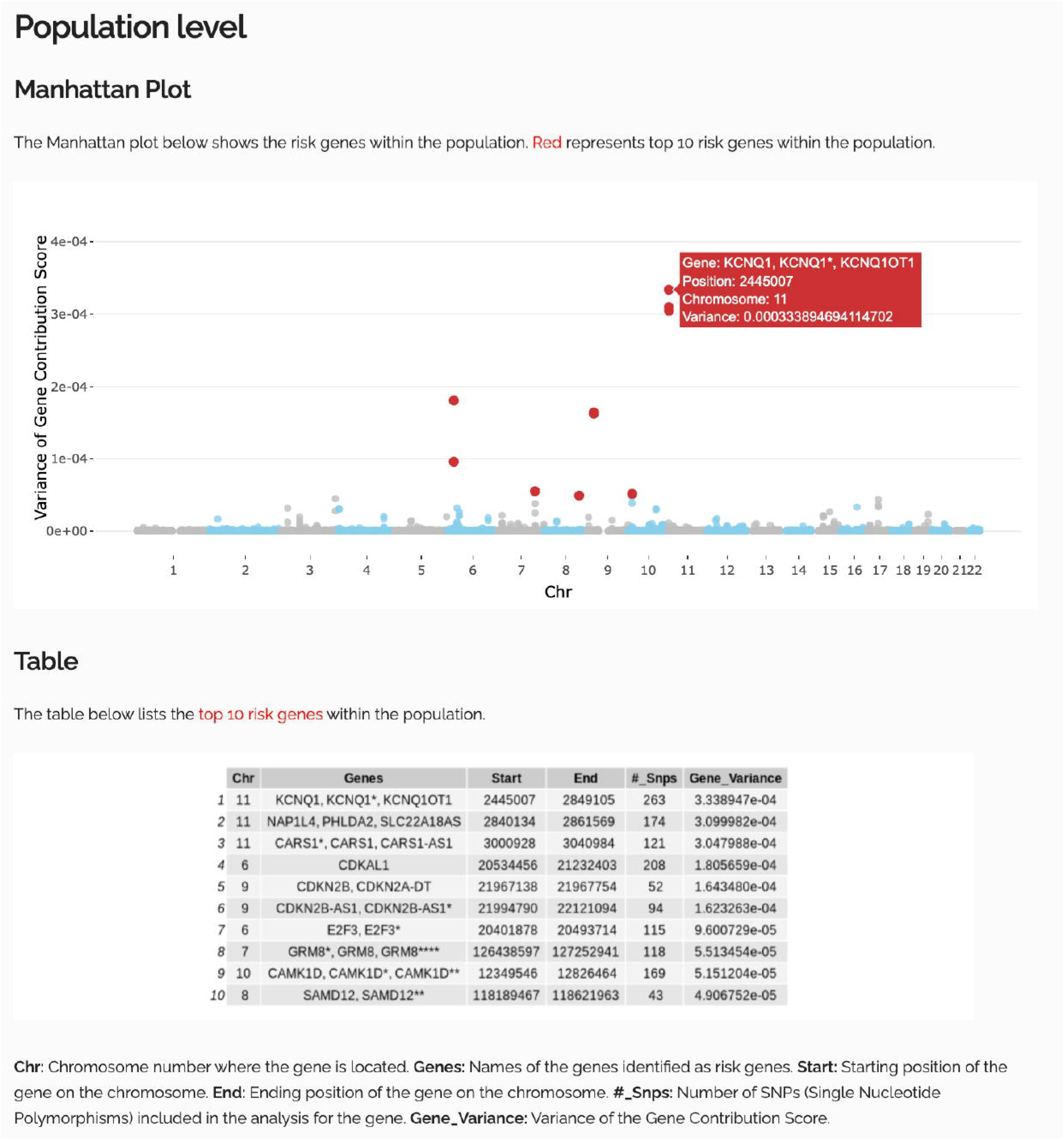
Contribution of genes to Type 2 Diabetes PRS in the Asian population from the 1000 Genomes Project. **Manhattan plot**: Variance in gene contribution scores across chromosomes. Higher variance indicates greater significance as a risk factor, with notable genes highlighted. **Table**: Significant genes, including chromosomes (chr), gene names, start and end positions, number of SNPs included in mapped genes, and variance in gene contribution scores.

### Visualizing the impact of genes and SNPs in an individual

To trust the PRS results and effectively communicate with patients, it is crucial to pinpoint which specific genes and SNPs contribute to the PRS of an individual. By visualizing the attributed values of gene and SNP contribution scores for each individual, we to increase the explainability of the PRS score. The example individual, HG00464, is obtained from the 1000 Genomes Project. Figure 3 shows the individual PRS within the population via a density plot, which shows the position of an individual relative to high or low genetic risk for type 2 diabetes. This individual has a high PRS, which is in the top 2 percent of the population.

**Figure 3.**
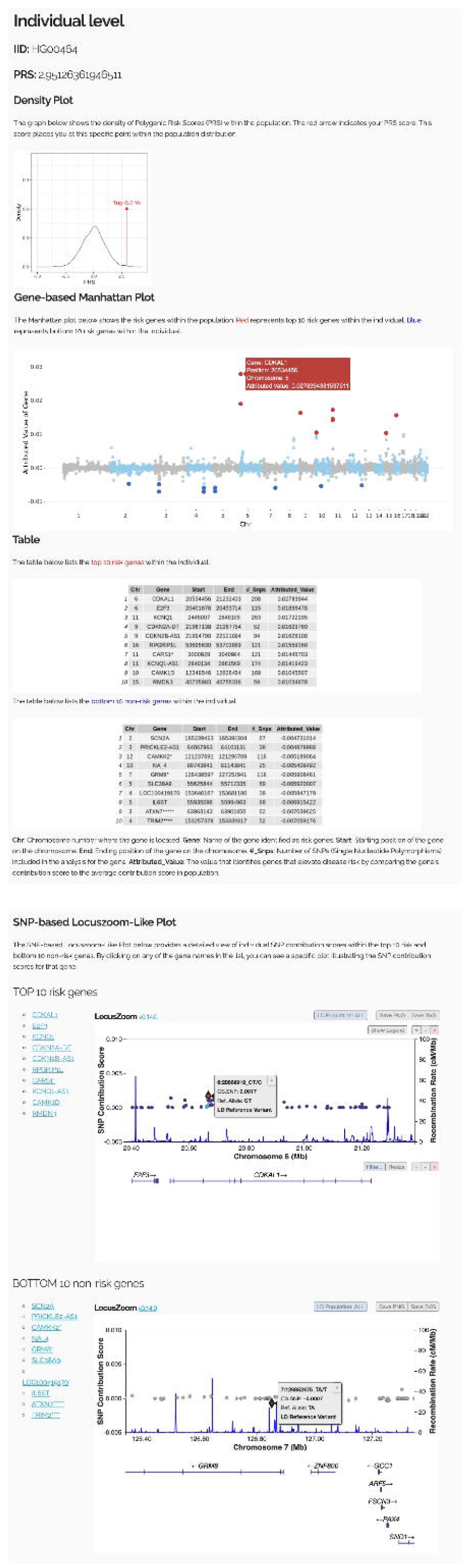
Effect of gene and SNP on an individual PRS from the 1000 Genome Project for type 2 diabetes. High PRS sample **IID**: HG00464. **Density plot**: PRS of the individual within the population distribution, indicating their genetic risk position. **Table:** Top 10 risk genes and bottom 10 nonrisk genes within the individual. **Manhattan plot**: Significant genes contributing to the PRS of the individual, with higher points representing greater impact. **LocusZoom-like plot**: Detailed SNP contributions for the top 10 risk genes and bottom 10 nonrisk genes

The figure includes a Manhattan plot and two tables to highlight specific genes that elevate or reduce the PRS. For example, the CDKAL1 gene, a well-known T2D-associated gene[20], had the highest attributed value. If this individual had the population average contribution value, the PRS score would be lower by 0.028. Additionally, the LocusZoom-like plot displays the detailed visualization of SNP contributions within the gene. For example, individual HG00464 carries 208 alleles in CDKAL1, which increases its PRS value. These figures enable the explanation of the PRS of an individual at both the gene level and SNP level, facilitating effective communication about their genetic risk.

On the other hand, in Supplementary Figure 1, the example individual, HG01816, has a significantly lower PRS than the population average does, indicating a lower genetic predisposition to type 2 diabetes. This figure also includes a Manhattan plot and table to emphasize specific genes that contribute minimally or even reduce the PRS. For example, KCNQ1, known as a diabetes-related gene[21], had the lowest attributed value. Additionally, the KCNQ1-AS1 gene, another gene associated with type 2 diabetes, had the second lowest attributed value, further contributing to the reduced PRS of the individual.

## Discussion

The XPRS tool enhances the interpretability of PRS by providing detailed contribution scores for individual genes and SNPs. By breaking down the PRS into its granular components, XPRS improves the explainability of these scores. Notably, identifying which factors contribute to genetic risk increases the reliability of predictions and strengthens user trust in the system. Furthermore, this approach will aid in effectively communicating genetic risk factors to key stakeholders, including clinicians and patients, ensuring that complex genetic information is more accessible and actionable in a clinical setting.

XPRS uses several visualization tools, including Manhattan plots and LocusZoom-like plots. At the population level, Manhattan plots were used to identify key genes contributing to disease susceptibility. For individual-level analysis, Manhattan plots show which genes contribute to risk, and LocusZoom-like plots provide a detailed view of specific SNPs that influence those genes. This visualization can effectively convey which SNPs or genes specifically influence the PRS of an individual.

While numerous models, such as P+T, LDPred, PRS-CS, BayesR, and MegaPRS, focus on constructing PRS by aggregating genetic variants to predict disease risk, the XPRS tool is distinct in its objective and functionality. Rather than constructing a PRS, XPRS is designed to interpret and explain existing PRS models. This allows XPRS to be applied across various PRS models, utilizing their scoring files to deliver insights at both the individual and population levels. By elucidating the underlying genetic factors contributing to the PRS of an individual, XPRS facilitates a more comprehensive understanding of genetic risk.

Despite its strengths, XPRS has certain limitations. Although XPRS is a web-based tool, privacy concerns require users to download and run the program on their own servers to ensure data security and compliance with privacy regulations. This requirement, while crucial for safeguarding sensitive information, may reduce the accessibility of the software. Additionally, since PRS relies on SNP-level predictions and XPRS interprets these predictions at the gene level, accurate SNP-to-gene mapping is essential for tool effectiveness. To increase the accuracy of SNP-to-gene mapping, we employed multiple strategies, including positional mapping, combined SNP-to-gene approaches, and methods based on GWAS p values or SNP heritability. However, SNP-to-gene mapping remains a complex and evolving field of research, which poses ongoing challenges for the accurate interpretation of genetic risk.

Explanation is crucial for the successful deployment of prediction models, yet currently, no explanation tools specifically designed for PRSs are available. Our research addresses this gap, facilitating the broader adoption of PRSs in clinical settings by providing the necessary interpretative tools to support informed decision-making.

## Key Points

- **Enhanced PRS Interpretability:** XPRS decomposes polygenic risk scores into gene/region and SNP contribution scores via Shapley additive explanations (SHAPs), providing clear insights into genetic factors influencing disease risk.
- **Comprehensive Visualization Tools:** The software offers multilevel visualizations, including Manhattan plots and LocusZoom-like plots, for population and individual analyses, facilitating effective communication between clinicians and patients.
- **Integration with Genomic Resources:** XPRS leverages genomic annotations such as refGene and cS2G mapping to ensure precise mapping of SNPs to their corresponding genes, increasing the reliability of PRS interpretations.
- **User-Friendly Platform:** Featuring an accessible cloud-based web interface, XPRS enables straightforward data input and interpretation.

## Acknowledgements

Not Applicable.

## Code Availability

The XPRS software is publicly available on GitHub at https://github.com/nayeonkim93/XPRS and can be accessed through our cloud-based web service at https://xprs.leelabsg.org/. XPRS integrates LocusZoom for visualization purposes and utilizes PLINK for genome-wide association analyses. LocusZoom is available at https://github.com/statgen/locuszoom and PLINK can be accessed at https://www.cog-genomics.org/plink/1.9/.

## Data Availability

SNPs are mapped to genes based on their genomic positions using the RefGene annotation from the UCSC Genome Browser (https://hgdownload.soe.ucsc.edu/goldenPath/hg38/database/). The cS2G file utilized in this process was obtained from Zenodo (https://zenodo.org/records/6354007).

## Supplementary Information

**Supplementary Figure 1.** Effect of gene and SNP in an individual PRS from the 1000 Genome Project for type 2 diabetes. Low PRS sample **IID**: HG01816. **Density plot**: PRS of the individual within the population distribution, indicating their genetic risk position. **Table:** Top 10 risk genes and bottom 10 nonrisk genes within the individual. **Manhattan plot**: Significant genes contributing to the PRS of the individual, with higher points representing greater impact. **LocusZoom-like plot**: Detailed SNP contributions for the top 10 risk genes and bottom 10 nonrisk genes

