## Supplementary figures and images for "XPRS: A Tool for Interpretable and Explainable Polygenic Risk Score"

### Supplementary Figure 1

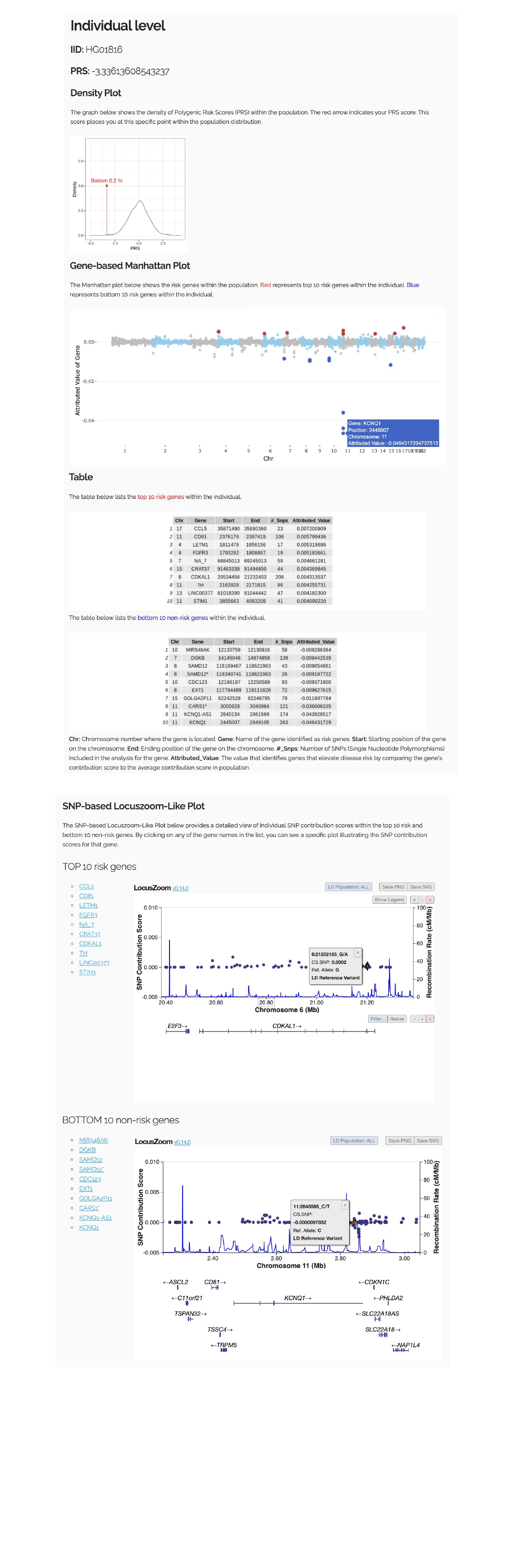
